# Real-time forecasting of COVID-19-related hospital strain in France using a non-Markovian mechanistic model

**DOI:** 10.1101/2023.02.21.23286228

**Authors:** Alexander Massey, Corentin Boennec, Claudia Ximena Restrepo-Ortiz, Christophe Blanchet, Samuel Alizon, Mircea T. Sofonea

## Abstract

**Background:** Projects such as the European Covid-19 Forecast Hub publish forecasts on the national level for new deaths, new cases, and hospital admissions, but not direct measurements of hospital strain like critical care bed occupancy at the sub-national level, which is of particular interest to health professionals for planning purposes.

**Methods:** We present a sub-national French framework for forecasting hospital strain based on a non-Markovian compartmental model, its associated online visualisation tool and a retrospective evaluation of the real-time forecasts it provided from January to December 2021 by comparing to three standard statistical forecasting methods (auto-regression, exponential smoothing, and ARIMA).

**Results:** For anticipating risk of critical care unit overload, our model performed worse than pure statistical methods at the one- and two-week horizons, but had better point forecasts at the four-week horizon for 8 of the 13 regions considered. Our model also suffered from over-confidence with respect to its prediction intervals.

**Conclusions:** Online visualisation tools and consideration of how metrics can be affected by distortion from non-pharmaceutical government interventions are essential in the assessment of forecasting models for hospital strain.

**What is already known on this topic:**

- The US and European Covid-19 Forecast Hubs do not provide more direct measurements of hospital strain like critical care bed occupancy at the sub-national level, which was essential for the provisioning of healthcare resources during the COVID-19 pandemic.
- In France, statistical modelling approaches have been proposed to anticipate hospital stain at the sub-national level but are limited by a two-week forecast horizon.

**What this study adds:**

- We present a sub-national French modelling framework and online application for anticipating hospital strain at the four-week horizon that can account for abrupt changes in key epidemiological parameters.
- It was the only publicly available real-time non-Markovian mechanistic model for the French epidemic when implemented in January 2021 and, to our knowledge, it still was at the time it stopped in early 2022.

**How this study might affect research, practice or policy:**

- Further adaptations of this surveillance system can serve as an anticipation tool for hospital strain across sub-national localities to aid in the prevention of short-noticed ward closures and patient transfers.

## INTRODUCTION

The COVID-19 pandemic emphasised that policymakers need access to accurate forecasts of key epidemiological indicators to mitigate strain on hospital services and reduce preventable deaths [1,2]. This led to international projects tasked with creating centralised repositories for COVID-19 forecasts pertaining to the United States [3], Germany/Poland [4], and Europe [5]. However, policies implemented at the local level tend to outperform uniform national policies [6]. Unfortunately, in contrast to their counterparts, the European COVID-19 Forecast Hub only considers the national level and countries like France had to rely on other sources to optimise their local to national healthcare system management.

Of particular interest is intensive care unit (ICU) occupancy, one of the most direct indicators of hospital strain, which is predicted using either statistical or mechanistic models. Statistical models use correlations in previously observed data to explain model structure that can be separated from noise and extrapolated to the future. In time series analysis, they can benefit from the addition of adequate predictor variables. For example, in France [7] proposed an ensemble model approach that combined several statistical models (including machine learning) to utilise predictors identified from available epidemiological, mobility, and meteorological data to make 14-day forecasts for ICU admissions and ICU occupancy by French region. Their ensemble was effective in the short-term but had more difficulty predicting beyond the lag (typically at most two weeks) between their predictors (e.g. positive antigen testing) and the subsequent hospitalisation events.

Mechanistic models are explicitly based on a simplified version of the underlying epidemiological process [8]. The most popular is the compartmental model, which typically involves separating a population into distinct sub-populations (e.g. susceptible, infected, and ‘removed’ individuals in the SIR model) and inferring the transition dynamics between these compartments. This can be done based on assumptions regarding the biology of the pathogen or via optimisation approaches using observed data, e.g. hospital admissions, to make inferences regarding partially or unobserved factors such as daily infections [9]. The biological assumptions simplify the causal relationships between a pathogen’s infectivity, pathogenicity, and lethality so that long-term forecasts can be produced. These models provide us with a mechanistic understanding of the epidemic process and can help to anticipate planned changes such as lockdowns or increases in vaccination coverage. A limitation of compartmental models is that they typically require large, idealised populations for best results [10] and can have lower predictive performance [11] depending on the time scale. However, as shown in [12], a non-Markovian discrete-time compartmental model may have the potential to capture the dynamics up to 5 weeks on average (although this also reflects the epidemiological relevance of the underlying assumptions made).

To facilitate COVID-19 monitoring in France, we developed COVIDici: a mechanistic transmission model that accurately captures the hospital and mortality dynamics from the French epidemic. Model results were publicly communicated via a web dashboard (https://cloudapps.france-bioinformatique.fr/covidici/), which provided real-time visualisations of the French epidemic at the national, regional, and departmental levels. COVIDici was updated daily using databases published by the national public health agency, Santé Publique France [13] until 2022 when it was halted after the emergence of the Omicron variant [14] led to decreased interest in epidemic forecasting by French authorities, partly driven by the belief that Omicron BA.1 would represent our way out of the pandemic and the last wave [15].

Here, we briefly summarise the structure of the underlying compartmental model, the statistical procedure for the parameter inference and describe communication via the web dashboard. We present an evaluation of COVIDici’s forecasts for ICU occupancy up to the four-week horizon at the regional and national levels using standard metrics for continuous variables as well as a binarized version representing ICU overload to focus on model performance in anticipating wave peaks. Standard statistical forecasting methods (auto-regression, exponential smoothing, and ARIMA) are included as baseline models. Finally, we discuss perspectives for COVID-19 epidemic modelling in the context of decreased surveillance.

## METHODS

This section presents a summary of COVIDici and a retrospective evaluation. COVIDici is based on a pre-existing discrete-time deterministic age-structured transmission model tailored to capture the dynamics of the hospital indicators of the epidemic in France, which was modified to include vaccination. Derivations of the flows and notational details are provided in [16]. Further technical details regarding the implementation of COVIDici as well as all the baseline models is accessible via the Supplementary Materials. The scripts and data used to perform the analysis and generate this manuscript are available on GitLab (https://gitlab.in2p3.fr/ete/covidici_public) and archived in Zenodo [17].

### The model

The structure of the model, shown in Figure 1, describes the flows between susceptible or infected people in the general community, hospitalised people, and individuals that no longer contribute to the epidemic due to recovery with full immunity or death. Stratifying by age, where the index denotes the individual age class, each box represents a group of individuals who share the same clinical kinetics and who contribute equally to the epidemic dynamics. Most susceptible individuals in will pass to the non-critically ill compartment, denoted with rate proportional to (for the vaccinated), for to days (for the vaccinated). A small fraction proportional to (for the vaccinated), which increases with age [18], of infected individuals reaches the critical infection compartment,, meaning they will eventually be hospitalised in critical care wards for to days or they will be in hospice care () for to days where they end up in with probability. Critically ill individuals move to with a rate proportional to and then die at a rate proportional to. However, if they enter they are assumed to have full immunity in the future., and reflect the expected distribution of days for individuals to exit, and respectively.

**Figure 1.**
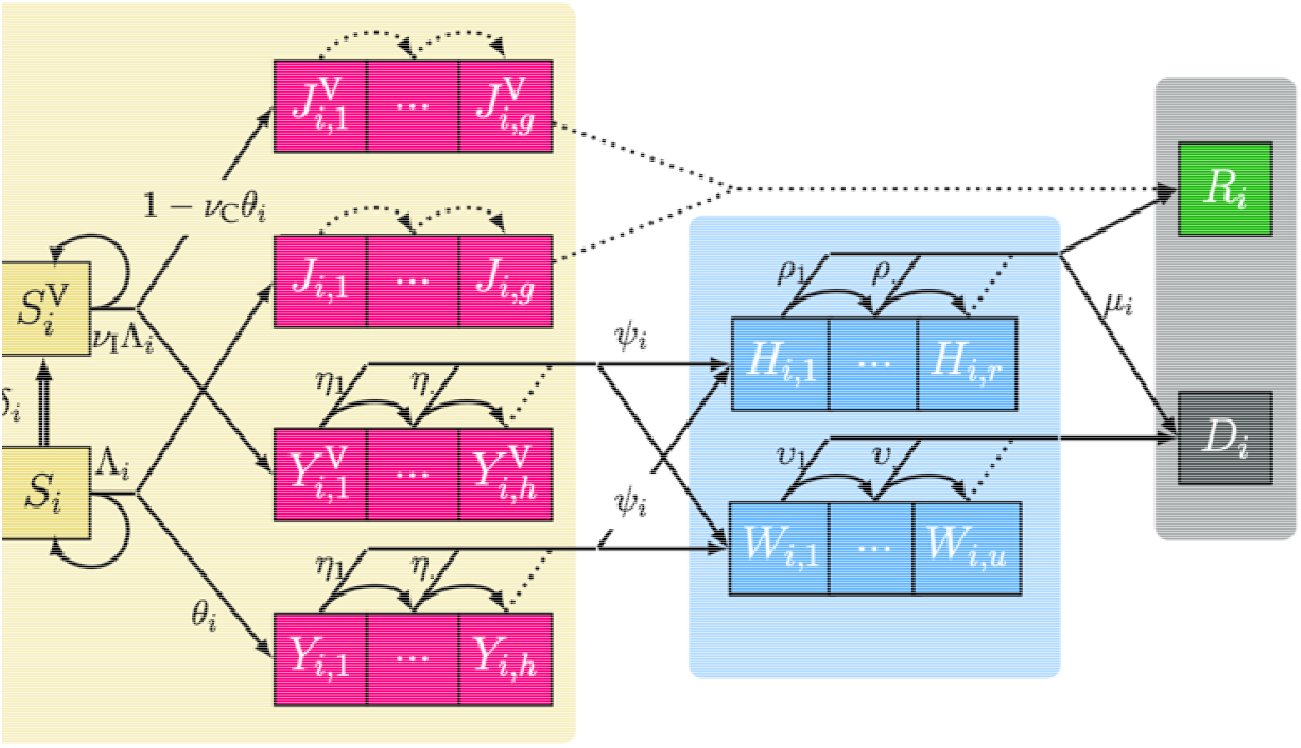
Structure of the underlying COVID-19 epidemic discrete time model with vaccination. The three shaded areas represent the general population (left), the hopitalised population (center) and individuals removed by either recovery or death (right).,, and are the maximum number of days possible to remain in a compartment represented by contiguous boxes. Individuals in are susceptible, are non-critically infected, are infected and will eventually require hospitalisation, are hospitalised in a critical care bed, are in hospice care, are recovered and are deceased., and denote vaccinated compartments. is the force of infection. in the vaccination rate.,, and are transition rates between compartments which may or may not be reduced by factors and as a result of vaccination. Arrows between boxes show the daily flow of individuals between compartments where dotted arrows occur with probability 1. For the sake of simplicity, only one age group (denoted using the subscript) is depicted here. Only one of the two complementary probabilities is shown for each bifurcating transition.

Aged care facilities (i.e. nursing homes) are ignored in this model because of their differences in epidemiology and hospitalisation rates compared to according to vaccination status prior to hospitalisation, with vaccinated compartments denoted by the exponent ^V^. is the anticipated vaccination rate for age class.

The most important parameter of the model is the rate at which susceptible hosts become infected and depends on the force of infection for a given time, defined as

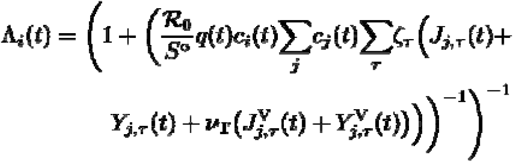

where is the basic reproduction number of the Wuhan strain of SARS-CoV-2 in France, is the French population size, and is a piecewise constant factor updating the reproduction number following variant replacement. and denote the per capita contact ratio of susceptible hosts (of age class) and infected hosts (of age class) respectively, and are set to for pre-pandemic values contact rates (note that this formulation assumes proportionate mixing between age classes). The index denotes the time in days since the beginning of the infection and allows us to vary host infectivity over the duration of the infection. For this, is the probability mass (i.e.∼the relative contribution) of the -th day of the infection regarding the generation time, for which we use the empirical serial interval distribution proposed by [19]. Thus, the model is non-Markovian in the sense that new infections do not only depend on the compartmental states from the previous day. Finally, the ratio captures the vaccine effectiveness against secondary transmission (i.e. the contagiousness decrease of breakthrough infections) [20].

### Modelling vaccination

The French national vaccination campaign started in late December 2020. To incorporate the vaccine rollout into the model, we explicitly assumed that the vaccines partially prevent infection and critical COVID-19 by reducing the force of infection experienced by all vaccinated susceptible hosts and the probability of being critically ill upon infection, set vaccine coverage in each age class in the model using the official VAC-SI database [21].

Future vaccination rates were predicted using a linear regression for each age group trained on the previous 3 weeks of vaccination data. We assumed that vaccination begins with the older age classes and that all age classes have an arbitrary vaccine coverage threshold of 90%. If the coverage for an age class was ever over this threshold, the doses planned for this age class were redistributed to the next oldest age class.

To avoid the inflation of the number of parameters, we assumed that full vaccination only required a single dose. Another simplifying assumption is that the vaccine is instantaneously effective, with an assumed permanent reduction in severe infection forms of 90% (*ν*_*C*_ = 0.1) and an 80% contagiousness reduction (*ν*_*T*_ = 0.2), which is an optimistic estimate [20].

## Calculation

Parameters were inferred under the Bayesian framework by running a Monte Carlo Markov Chain algorithm over a set of 12,000 realisations of the model. The last 2,000 iterations were used to generate the median and the 95% credibility interval for the parameters and 95% forecasting range for the time series, assuming a Poisson likelihood for daily ICU admissions. Except for the reproduction number ℛ_O_, the expectation and the variance of the infection-to-hospitalisation delay, which were inferred at the nationwide level only, all the parameters were independently fitted for each sub-national administrative division. Details on the inferred parameters, their prior values and distributions are provided in [16].

We expected some of these parameters to change over time due to virus evolution, e.g. the increased transmissibility of the Alpha [22] and then of the Delta variant [23], but also to public health interventions (e.g. lockdowns, curfews, limitations on businesses, etc.), social factors (e.g. school holidays), improvement in COVID-19 patient care, and variation in patient profiles. To account for this, we allowed for some parameters to be time-dependent by partitioning the time since the beginning of the epidemic and allowing each period to be associated with its own parameter set.

Parameter estimations were optimised based on the daily COVID-19-related critical care admissions from the COVID-19 hospital activity database (SI-VIC) [13]. In France, critically ill patients can be hospitalised either in intensive care units, continuous care units, or acute care units, the three forming the critical care capacities [24]. For simplicity here, ICU refers to the wider category of critical care beds, as provided by SI-VIC. Furthermore, we assume the age distribution between localities to be fixed and based on the official demographics data [25].

### Communication

An automated cluster computing workflow refit the COVIDici model using daily updates of hospital, vaccination and testing data downloaded from the SI-VIC database, allowing a Shiny web application (see Figure 2 for screenshot, link in Introduction or [17] source code) to communicate real-time results to the public. The original 2021 production version permitted users to visualise the combined past and future model fit by national, regional or departmental administrative unit for multiple epidemiological parameters, including ICU admissions, ICU occupancy, mortality (cumulative and daily), temporal reproductive number (*R*(*t*)), infections (cumulative, daily and current), vaccination coverage and incidence for positive tests.

**Figure 2.**
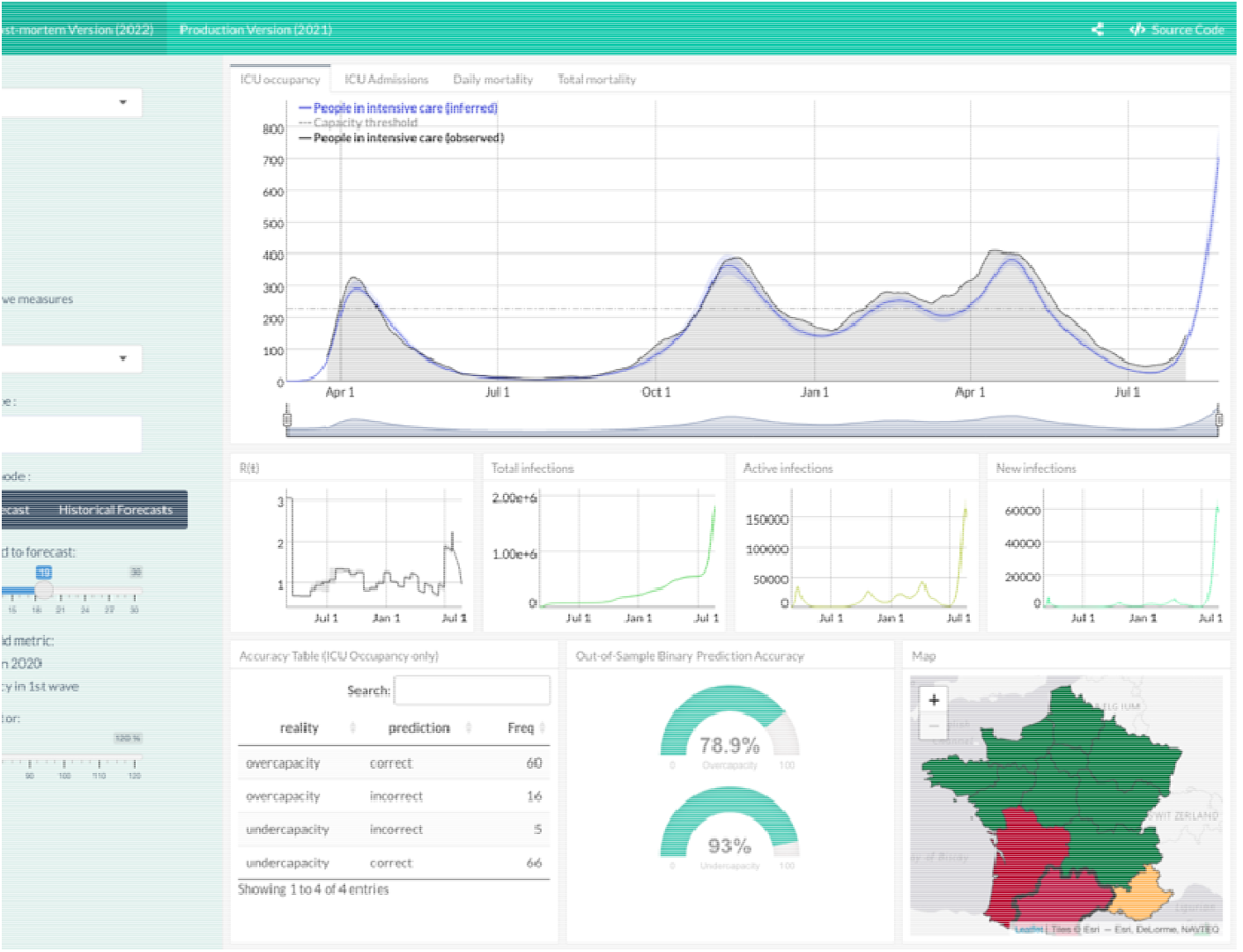
Screenshot of the COVIDici application showing model fit for daily ICU occupancy in the Occitanie region.

In 2022, a post-mortem version of the interface was deployed to allow for retrospective inspection of past forecasts with respect to a historical reference date. This version allows visualisation of all forecasts occurring prior to the reference date and includes basic evaluation metrics based on ICU overload (i.e. binarized ICU occupancy) and a colour-coded heatmap of hospital strain for varying forecast lengths and arbitrary saturation thresholds.

### Retrospective evaluation

Our assessment is based on original forecasts made by COVIDici between January 30, 2021 and the first detected Omicron case in France (i.e. December 2, 2021), taken on a weekly basis to match with evaluation frameworks of the European and US Covid-19 Forecast Hubs. We focus here on ICU occupancy and only consider the regional and national levels while emphasising that many of the results are equally as valid on the departmental level, especially when they contain major urban areas.

The following baseline models were evaluated using a rolling forecasting origin [26] starting on August 2, 2020:

- **ETS+ARIMA** is an ensemble of an ARIMA and an exponential smoothing (ETS) model fit using automated defaults in the *fable* package in R. It uses a log transformation of the rolling 7-day average of the ICU occupancy using only the data available for COVIDici to make its original forecast for that reference date.
- **AR-Lasso** is an auto-regressive (AR) machine learning type model that does not require stationarity and was implemented using the *caretForecast* package. Lagged values from the previous 21 days are selected using the Least Absolute Shrinkage and Selection Operator (LASSO) [27] tuned using time-series cross-validation as implemented in [28] to prevent data leakage and reduce overfitting. Prediction intervals were calculated using a bootstrap of the one-step ahead residuals from the training fit.
- **Naive** is a special case of an AR-1 implemented using the *fable* package. The point forecast is simply the last observed value and is optimal if the time series is a random walk.

### Standard metrics

We define “standard metrics” as those recommended by the US and European COVID-19 Forecast Hubs using the same 23 quantiles of the forecast distribution. Thus, point forecasts (based on the median) are evaluated with the absolute error (AE), individual prediction levels with the empirical coverage rate (ECR), and the forecast distributions with the weighted interval score (WIS). The WIS is a proper scoring rule that generalises the absolute error and gives penalties for interval spread as well as for over- and under-prediction. Formally speaking, for the true value of the target variable, we have a forecast distribution with median that contains a set of prediction limits are the and quantiles of the. WIS is defined as the following weighted sum:

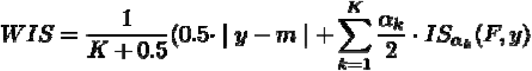

where for a single interval, the interval score is computed as

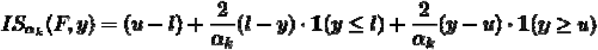

with being an indicator function and the nominal coverage of interval. Note that if only the median is included (i.e.) then the WIS simplifies to the AE. Furthermore, as the number of equally spaced intervals increases, the WIS converges to the Continuous Ranked Probability Score. We refer readers to [29] for a deeper technical explanation).

All three standard metrics (AE, ECR, WIS) were calculated using the *scoringutils* package [30]. We used the package’s default summary function (i.e. the mean) when aggregating over geographic units. However, we use the median when aggregating over time because the mean tended to give distorted results due to outlier errors that are common during the peaks of epidemiological waves (see Figure 3A).

**Figure 3.**
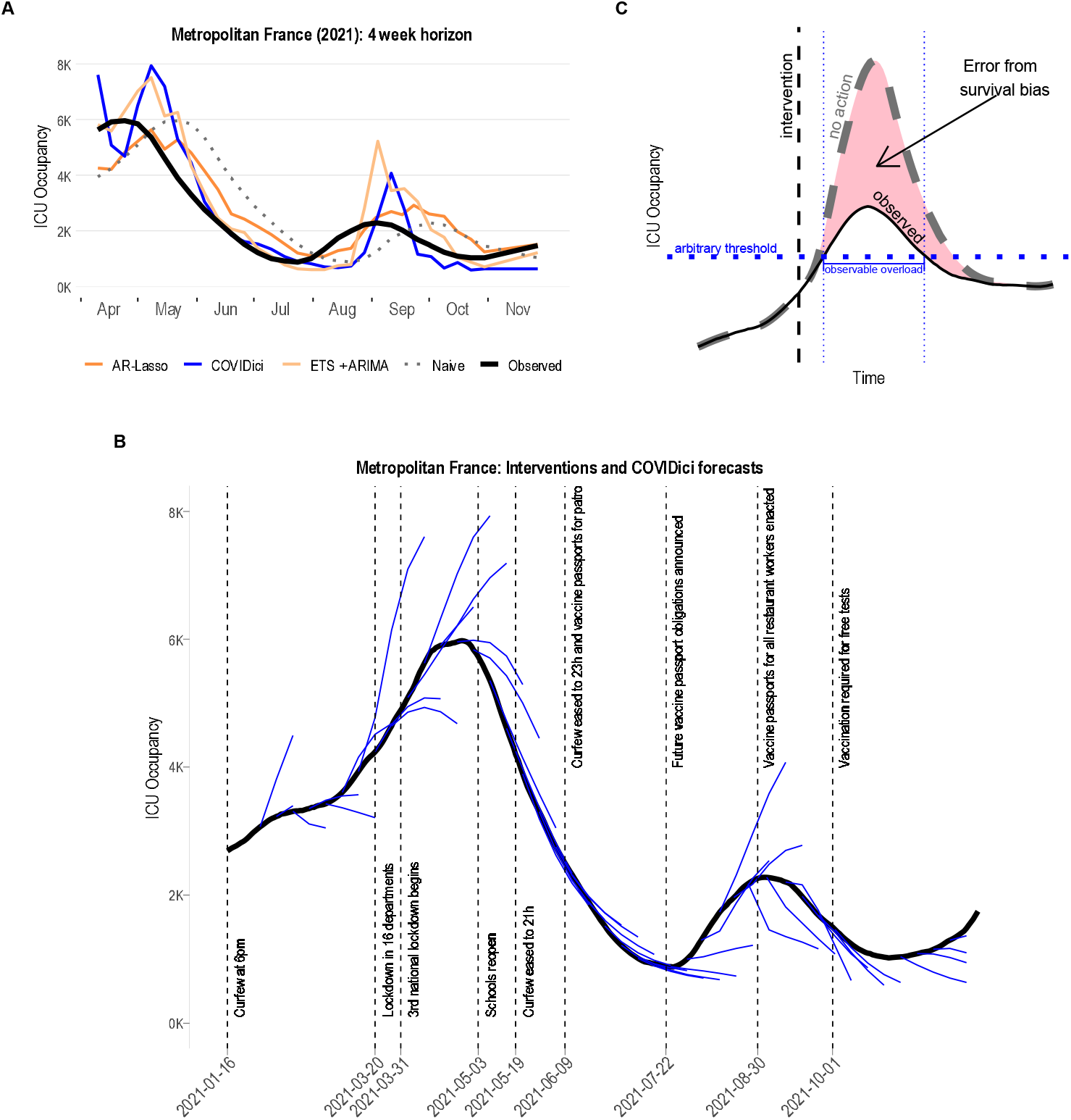
Qualitative inspection of COVIDici forecasts. A) All forecasters at the 4-week horizon plotted with the observed ICU occupancy for metropolitan France. B) Overlay of national forecasts of ICU occupancy produced by COVIDici with list of governmental interventions issued at the national level. C) Schematic representation of the survivor bias that may occur after a restrictive governmental intervention. The dashed curve is the ICU occupancy that would have occurred if the intervention did not happen. The solid curve is the ICU occupancy that we observed happen because the intervention did occur.

Summarising AE and WIS with the median have the drawback that it is more likely to reflect forecaster performance between waves rather than its ability to anticipate peaks, which is arguably the more important objective. Furthermore, AE and WIS tend to harshly punish outlier errors during wave peaks which can at least be partially explained by a survivor bias that occurs every time a public health policy is implemented (see Figure 3B for non-exhaustive list), as well as spontaneous behavioural change. As illustrated by Figure 3C, this bias (i.e. the shaded area between the curves) corresponds to the difference between what would happen in absence of intervention (dashed curve) and what eventually is observed (the solid curve). While the magnitude of this bias at the peaks is counterfactual and subject to debate, several studies have indicated that even mild non-pharmaceutical interventions can have similar effects on curbing the spread of the virus compared to more severe ones [31,32].

### Binary metrics

To fairly evaluate performance during wave peaks, we consider ICU overload (binarized ICU occupancy), which we expect to be more robust against over-predictions. This requires introducing arbitrary capacity thresholds which we define as the percentage of the ICU occupancy observed in the geographical unit in the first wave in 2020. For point forecasts, we consider the proportion of incorrect forecasts of an outcome given that outcome was observed. Following the convention that lower scores are better, we define:

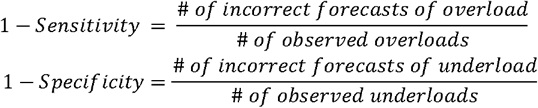

For forecast distributions of ICU overload, we use the Brier score, which is the mean squared error of the binary overload outcome (i.e. 0 or 1) and the mass of the prediction interval above the arbitrary threshold. Formally, this is defined as:

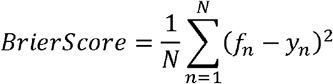

Where *f*_*n*_ is the predicted probability of overload event *y*_*n*_ ∈ {0,1} with *n* = 1,… *N* denoting all the events in the scope of the evaluation. The predicted probability *f*_n_ can be approximately calculated as

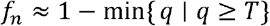

Where is the collection of quantiles of the forecast distribution and the arbitrary threshold. We present binary metrics separately for periods of observed overload and underload because anticipation of overload is widely considered more important in a hospitalisation surveillance system.

## RESULTS

Qualitatively, all four forecasters performed reasonably well at the four-week horizon (see Figure 3A) although COVIDici and ETS+ARIMA tended to over-predict more at the top of the waves. All forecasters experienced a delay in anticipating upcoming waves, which was more pronounced for COVIDici and the naive model. Figure 3B shows qualitative evidence that COVIDici performs better on the trailing side of each wave than the leading side.

Standard metrics for ICU occupancy are contained in Figure 4. Figure 4A shows the mean WIS (scaled by the naive model) across all geographic units over time. Large misses near wave peaks are evident for COVIDici and ETS+ARIMA but not AR-Lasso, which appears to be more consistent. This narrative is reversed in Figure 4B which depicts the median AE and WIS by region. COVIDici clearly shows improvement relative to the other baselines at longer horizons and had the lowest AE at the four-week horizon for 8 of the 13 regions. These improvements were not present when evaluating the forecast distribution using WIS. This is likely explained by Figure 4C, where we see strong evidence that the prediction intervals for COVIDici were far too narrow. AR-Lasso was somewhat better calibrated and ETS+ARIMA had near optimal calibration of predictions intervals nationally as well as in several regions.

**Figure 4.**
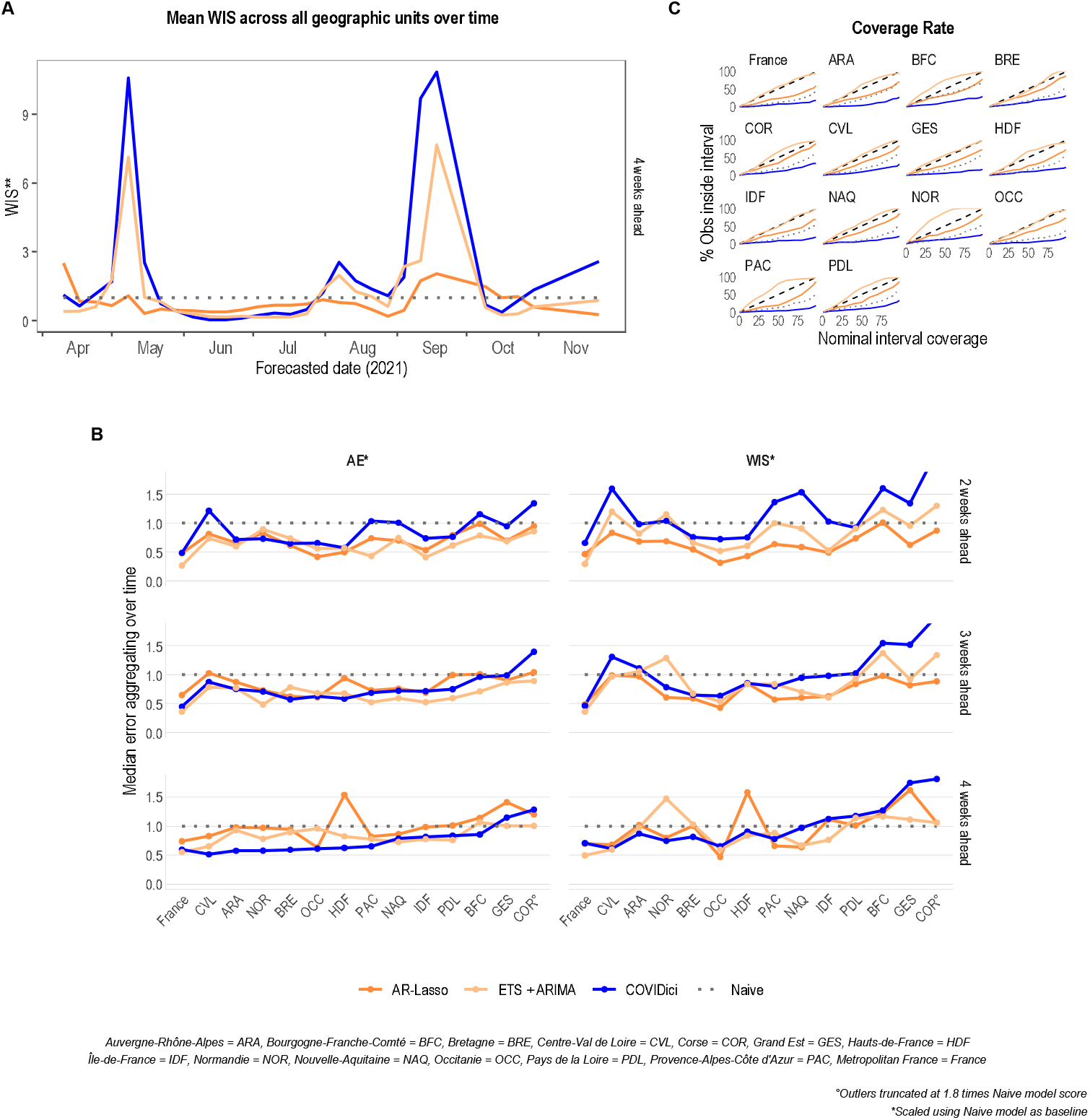
Standard evaluation metrics for ICU occupancy. A) Mean weighted interval score (WIS) of all geographic units over time for the four-week forecast horizon. B) Absolute error (AE) and WIS per geographic unit by two-to four-week horizons. Aggregation over time uses the median here because the mean is highly sensitive to outlier errors during the waves. C) Empirical coverage rate across all geographic units and forecast horizons (dashed line is optimal).

Binarized metrics for ICU overload are shown in Figure 5. All binary metrics show that COVIDici consistently improves relative to the baselines at longer horizons. Figures 5A and 5B show all evaluations of point and distribution metrics for ICU overload at varying capacity thresholds. AR-Lasso is clearly not suitable for anticipating ICU overload at the four-week horizon where it only performed comparably to the naive baseline while ETS+ARIMA had slightly fewer incorrect forecasts of overload compared to COVIDici but slightly more when predicting underload. ETS+ARIMA and COVIDici were also the top two choices at the four-week horizon in Figure 5B where the former had a consistent edge likely due to the overly narrow predictions intervals of COVIDici. Figure 5C breaks these metrics down by region which supports similar conclusions.

**Figure 5.**
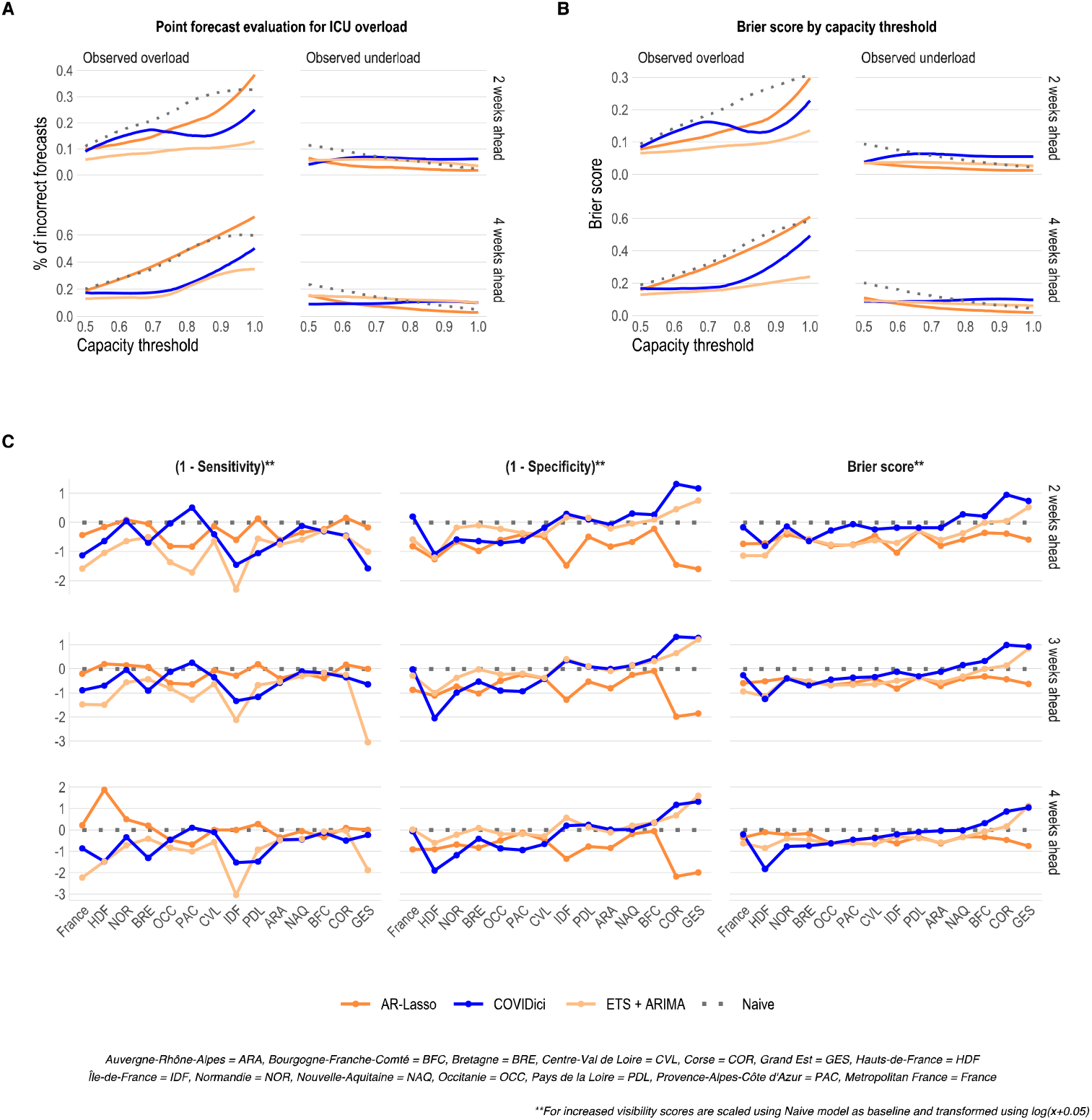
Binary metrics for ICU occupancy overload. A) Smoothed regression of point forecast scores by threshold after aggregating over all geographic units and time. B) Smoothed regression of distributional forecast scores by threshold after aggregating over all geographic units and time. C) All binary metrics by region aggregating across all capacity thresholds and time. The 51 capacity thresholds span between 0.5 and 1 and represent the percentage of each respective geographic unit’s peak ICU occupancy observed in the first wave in 2020.

## DISCUSSION

### Modelling and forecasting

Mathematical modelling was vital to understanding the COVID-19 epidemic in real-time as it unfolded. In France, it led to the anticipation of hospital dynamics in the first epidemic wave [33] and the emergence of the Delta variant[23]. However, most of these studies were performed at a national level and/or at a single time point, which made their impact limited from a public health point of view. To our knowledge, there were only two continuously running forecasting models in France in 2021 at the subnational level: COVIDici and an ensemble of statistical models implemented by the Pasteur Institute [7]. These two models were built on two types of approaches each with different strengths and limitations. For the latter this meant reasonable forecasting accuracy but only for relatively short forecast horizons.

In addition to real-time sub-national forecasting, a key feature of COVIDici was to offer visualisation of numerous unobservable indicators that can only be inferred through an underlying mechanistic model (e.g. the estimate of all active infections, reproductive number, etc.). This is an asset from a popularisation point of view, but it raises issues because estimates can be strongly biased in areas with low population density.

Furthermore, it is important to distinguish between two types of forecasts. Some, like COVIDici, attempt to capture what will happen if transmission remains identical, i.e. in absence of intervention or behavioural change. Others, especially the ones built on machine learning, try to factor in these changes to forecast what will eventually happen. In terms of guiding public health decisions such as the allocation of resources, the former seems more appropriate than the latter. However, this requires familiarising the audience with the well-known dilemma that we expect forecasts made under the assumption that nothing changes to be proven wrong by the observed data when they are too pessimistic. Otherwise, it indicates they were not considered when shaping the public health response.

### Perspectives

Many countries are decreasing their investment in epidemic surveillance, and some rely on statistical model forecasting with the inclusion of new predictors, such as that from wastewater data. However, there is still room for compartmental model forecasts like COVIDici that can rely on variables with a high level of sampling such as hospital admissions data.

Regarding SARS-CoV-2, future extensions of COVIDici would require updating the model to account more precisely for the diversity in immune protection among individuals, given the number of natural infections since the evolution of the Omicron variants [14]. However, existing non-Markovian models suggest that this is feasible [34].

Furthermore, in several ways COVIDici’s forecasting potential was under-exploited. It already incorporated variations in vaccine coverage but, thanks to its mechanistic nature, it could also readily include planned events such as school holidays or early predictors of variations in reproduction numbers such as temperature.

## Conclusion

Our benchmarking analysis revealed two major limitations for COVIDici. First, the prediction intervals were far too narrow which obfuscated any performative advantages that it had when considering distributional evaluation metrics (e.g. WIS and Brier score). Second, COVIDici exhibited relatively weak performance up to the two-week horizon to predict ICU occupancy compared to statistical modelling approaches. However, this was expected as COVIDici only uses hospital data to update its inference and there is a nearly two-week delay between infection and hospital admission [34].This decreased performance at the one- and two-week horizons relative to a benchmark is also consistent with findings regarding other mechanistic compartmental models predicting cumulative deaths at the national level in the United States [35].

The main strength of COVIDici was improved accuracy at the four-week horizon for point forecasts of ICU occupancy compared to statistical methods, especially during the trailing edge of waves. For anticipating wave peaks, COVIDici had one of the best overall performances with respect to the trade-off between the correct prediction rate of observed overload and observed underload on the four-week horizon. The baseline model based on machine learning (i.e. AR-Lasso) on the other hand failed to reasonably anticipate overload, despite relatively optimistic performance in terms of more standard metrics for continuous variables such as WIS. This should serve as a cautionary example for similar models that avoid large errors by avoiding large predictions at peaks of the waves. Systematically avoiding pessimistic predictions may improve the resulting evaluation score, but it also may complicate decision-making regarding non-pharmaceutical government interventions which is a common goal when forecasting hospital strain. To detect models exhibiting such undesirable behaviour, it seems reasonable to consider alternative evaluation metrics based on ICU overload (i.e. binarized ICU occupancy) especially when the evaluation period contains frequent interventions in the training sets.

## Data Availability

The model in this manuscript is based on publicly available data provided by Santé Publique France. The scripts and data used to perform the analysis and generate this manuscript are available on GitLab (https://gitlab.in2p3.fr/ete/covidici_public) and archived in Zenodo (doi:10.5281/zenodo.7641133). The companion web dashboard is hosted by France Bioinformatique (https://cloudapps.france-bioinformatique.fr/covidici/).

https://www.data.gouv.fr/fr/datasets/donnees-relatives-aux-personnes-vaccinees-contre-la-covid-19-1/

https://www.data.gouv.fr/fr/datasets/donnees-hospitalieres-relatives-a-lepidemie-de-covid-19/

https://gitlab.in2p3.fr/ete/covidici_public

https://cloudapps.france-bioinformatique.fr/covidici/

## ACKOWLEDGEMENTS

We acknowledge support from the MODCOV19 platform set up in March 2020 thanks to the National Institute of Mathematical Sciences and their Interactions (INSMI) and the CNRS (PaSSES grant) and the MODVAR project under the EMERGEN consortium. We also want to thank Santé publique France, the University of Montpellier, the South Green computational platform/IRD and the French Institute of Bioinformatics.

## Notes

### Competing Interest Statement

The authors have declared no competing interest.

### Funding Statement

This study was funded by the PaSSES grant under the MODCOV19 platform (INSMI and CRNS).

### Author Declarations

The study used ONLY openly available human data that were originally located at Santé Publique France.

### Summary of Updates

Minor corrections in Supplementary Materials where references to other journals to which submission was being considered are removed. More mathematical details in the Supplementary Materials were included in main text. Supplementary details is shortened. Limitations of the model were more explicitly addressed in abstract and conclusion.

