## Supplementary Materials for "Real-time forecasting of COVID-19-related hospital strain in France using a non-Markovian mechanistic model"

April 6, 2023

### Purpose

This document provides further details and mathematical formulations to sections as presented in the publication with the aforementioned title. Sections in the supplement generally correspond to the same section titles of the main article.

### Calculation

Summary values of the main simulated parameters of interest (e.g. ICU admissions, ICU occupancy, etc.) were originally saved using only the mean and the two quantiles $q_{2.5}$ and $q_{97.5}$. However, it has become commonplace in forecast hub projects (i.e. [1]) to consider as many as 23 quantiles in order to assess the entire predictive distribution at a small granular level. For the retrospective evaluation, this necessitated the imputation of missing quantiles that were not saved under the standard COVIDici workflow in order to conduct a similar comparison. As a result, we fitted a skewed normal distribution when the point forecast value was greater than 6 (daily events) or a log-normal distribution when the point was less than 6 (i.e. close to zero) using quantile matching. Code and documentation for this post-processing step is available in our public git repository [2].

### Communication

The first step in the updating phase was the upload of the vaccination, critical care, and mortality data every evening after it had been updated by Santé Publique France [3]. During the night, a job was assigned to each territory that creates a workspace on a node of the cluster, which contained all the data and scripts necessary to derive all the parameters and created time series of each type of observable data on the COVIDici application (ICU admissions, ICU occupancy, etc.). The outputs were generated as CSV files and repatriated to the same cluster in a git project. This was followed by an update of the git project on the project server, which contained the code running the interface and all the data used by it. In the morning, the Shiny application was updated by the Institut Français de Bioinformatique (IFB) and made available at: `https://cloudapps.france-bioinformatique.fr/covidici/`.

### Retrosective evaluation

#### Criteria for inclusion

Although COVIDici updated its model and public app interface daily, we adopted the format used by the European COVID-19 Forecast Hub to construct our retrospective evaluation for consistency. Therefore, we recovered historical git submissions made on Sundays (or at the latest Mondays in two cases) between January 30, 2021, to December 2, 2021, the day of the first detected infection from the Omicron variant in France. We then extracted weekly forecasts for the 1- to 4-week horizons which corresponded to target forecast dates always falling on subsequent Saturdays. The last day of available hospital data used to update COVIDici usually corresponded to the day before the submission and as early as the previous Thursday, depending on the data publication delay for a given region. A few of the weeks had no submissions because of server maintenance to the cluster. These delays, as well as the occasional revision of official estimates, could not be avoided and represent some practical aspects of real-time forecasting.

Original estimates were produced and presented on a daily basis using rolling averages to smooth any weekly effect. Evaluating forecasts for hospital demand on a daily basis is problematic because the actual number of hospital admissions on a given day tends to be systematically under-reported on weekends and over-reported during the beginning of the following work week. This data artefact, known as weekly seasonality, leads to a discrepancy between actual hospital demand (i.e. what we want to know but is unobservable) and the figures officially reported by state agencies. We presented our forecasts on a daily level because increased granularity is more useful from a planning perspective, including the potential impact of non-pharmaceutical interventions known to be implemented on a given day. However, forecasts were evaluated on a weekly basis to limit the weekly seasonality and because collaborative forecasting projects such as the European and US Covid-19 Forecast Hubs already use weekly evaluations.

### Target variable

Since COVIDici is based on a mechanistic model, it provided daily updates of coherent estimates for many epidemiological indicators of interest such as daily and cumulative mortality, daily and cumulative infections, current infections, ICU admissions, and temporal reproduction number at the French national, regional and departmental levels. For the evaluation, we only focus on forecasts of ICU occupancy because it is the most relevant indicator of hospital strain available in the SI-VIC database. Readers wishing to download this hospital data for themselves are encouraged to consult ./scripts/update_master.R in [2] to find the relevant URLs to download directly from SI-VIC.

### Baseline models

Baseline models were evaluated based on a rolling forecasting origin (with non-fixed window length) starting on August 2, 2020. It should be noted that several other baselines were originally considered, e.g. auto-regressive (AR) models using various boosting techniques for variable selection potentially with predictors derived from testing data, and are still available in the provided code [2]. Ultimately, since predictor variables themselves need to be forecasted in the future and the expected delay between positive testing is not longer than 2 weeks, the following three baseline models were selected in order to simplify the presentation of the results:

- **ETS+ARIMA** is an ensemble of an ARIMA and an exponential smoothing (ETS) model fit. It uses a log transformation of the rolling 7-day average of the ICU occupancy up until the last day of data available to COVIDici for each week matching the inclusion criteria. The ensemble was implemented using the fable package in R with the default settings to automatically identify model parameters for the 43 weeks of the evaluation period and 14 national and sub-national geographic areas. ARIMA requires that the time series is stationary which was automatically detected with package defaults using differencing. Prediction intervals were simulated by sampling from the residuals assuming a normal distribution. This baseline is consistent with classical statistical modelling approaches for time series without the inclusion of external covariates. Mathematical details and a full tutorial regarding the implementation of these models are available at [4]. Code implementing the ensembling procedure is available in ./scripts/classical_forecasts.R of the git repository.

- **AR-Lasso** is an AR machine learning type model implemented using the caretForecast package. This model uses a Box-Cox transformation to stabilise the variance and a linear regression for the past 21 lags as well as Fourier terms to account for potential weekly seasonality. It performs variable selection and regularisation using least absolute shrinkage and selection operator (Lasso) to optimise predictive performance. Formally, the Lasso coefficient vector, $\beta \in \mathbb{R}^p$, minimises the following loss function:

$$(Y - X\beta)^T(Y - X\beta) + \lambda|\beta|_1 \quad (1)$$

where $|\beta|_1 = \sum_{j=1}^{p} |\beta|_j$, $Y \in \mathbb{R}^n$ is the time series of the target variable (e.g. ICU occupancy), $X \in \mathbb{R}^{nxp}$ is the design matrix containing relevant lags of the target variable and Fourier terms for seasonality and $\lambda$ is the tuning parameter optimised using time series cross-validation as described by [4] with a 14-day cross-validation horizon. Predictions intervals were recursively simulated using a bootstrap of the one-step ahead residuals from the training set, summarised into predicted quantiles, and then smoothed with a 7-day rolling average to be compared to COVIDici. Thus the seasonality was accounted for in the model and then removed afterwards. This model is included mostly as a cautionary example of how certain evaluation metrics can be misleading. In practice, it avoids large errors by avoiding large predictions at peaks of the waves, which is an undesirable behaviour.

- **Naive** is a baseline and a special case of an AR-1 where the first lag is an offset. It was also implemented using the fable package. The point forecast is simply the last observed value and is optimal if the time series is a random walk (i.e. unpredictable white noise). Essentially this model is the best choice if the modeller believes that there is no exploitable structure in the time series to be used in a prediction. Note that the naive model will always have at least one (near) perfect prediction for a given forecast horizon towards the top of waves because it will catch the trailing side of the wave as the numbers inevitably decrease after the peak. This produces an artifact in evaluation metrics that are scaled using the naive model because this tends to occur at a time when other more reasonable forecasters overshoot the top of the wave producing particularly abnormally bad evaluation scores.

#### Limitations of standard metrics

Effectively evaluating forecasters at the wave peaks is further complicated by the fact that over-predictions might be (at least partially) explained by survivor bias that occurs every time a non-pharmaceutical government intervention is implemented. For example, in COVIDici, we predict the future outcome that would occur assuming no change in the transmission pattern (meaning no intervention and no spontaneous behavioural change), but for the evaluation we only observe the future outcome that occurred with the intervention.

Figure 1 illustrates this notion of survival bias. The dashed line is the counterfactual ICU occupancy that will hypothetically occur without intervention, i.e. what we want to estimate. The black line is the observed ICU occupancy that we observe (i.e. with potential interventions). The shaded area is the undeserved absolute error we would see due to survivor bias if we had a perfect prediction of the dashed line. Conversely, if we had a perfect prediction of the "observed" ICU occupancy, then our absolute error would be scored as zero despite the "true absolute error" being the shaded region. Furthermore, metrics related to the forecast distribution, i.e. the coverage rate and WIS, could also be substantially affected by this bias.

**Binary metrics**

To mitigate the aforementioned limitations, we introduce an additional evaluation metric that is robust against the negative effects of survival bias. We do this by evaluating forecasts with arbitrary thresholds and converting the target variable into a binary variable representing ICU overload and underload. To allow for comparability between geographical units with different population sizes, we defined the threshold as the percentage of the ICU occupancy observed in that geographical unit in the first wave in 2020.

One logical metric is correct prediction rate given that overload was observed, which is also known as the sensitivity. In Figure 1 we see that in areas of observable overload defined with respect to an arbitrary threshold, the dashed line that we want to predict will always predict overload as well under the very reasonable assumption that government interventions intended to reduce hospital strain do not increase it. This means that forecasters, such as COVIDici, attempting to predict the dashed line will not be unfairly penalised for forecasting what they are intended to.

Computing the specificity of our estimates, i.e. the correct prediction rate given that underload was

Figure 1 – **Schematic representation of the survivor bias that may occur after a restrictive governmental intervention.**

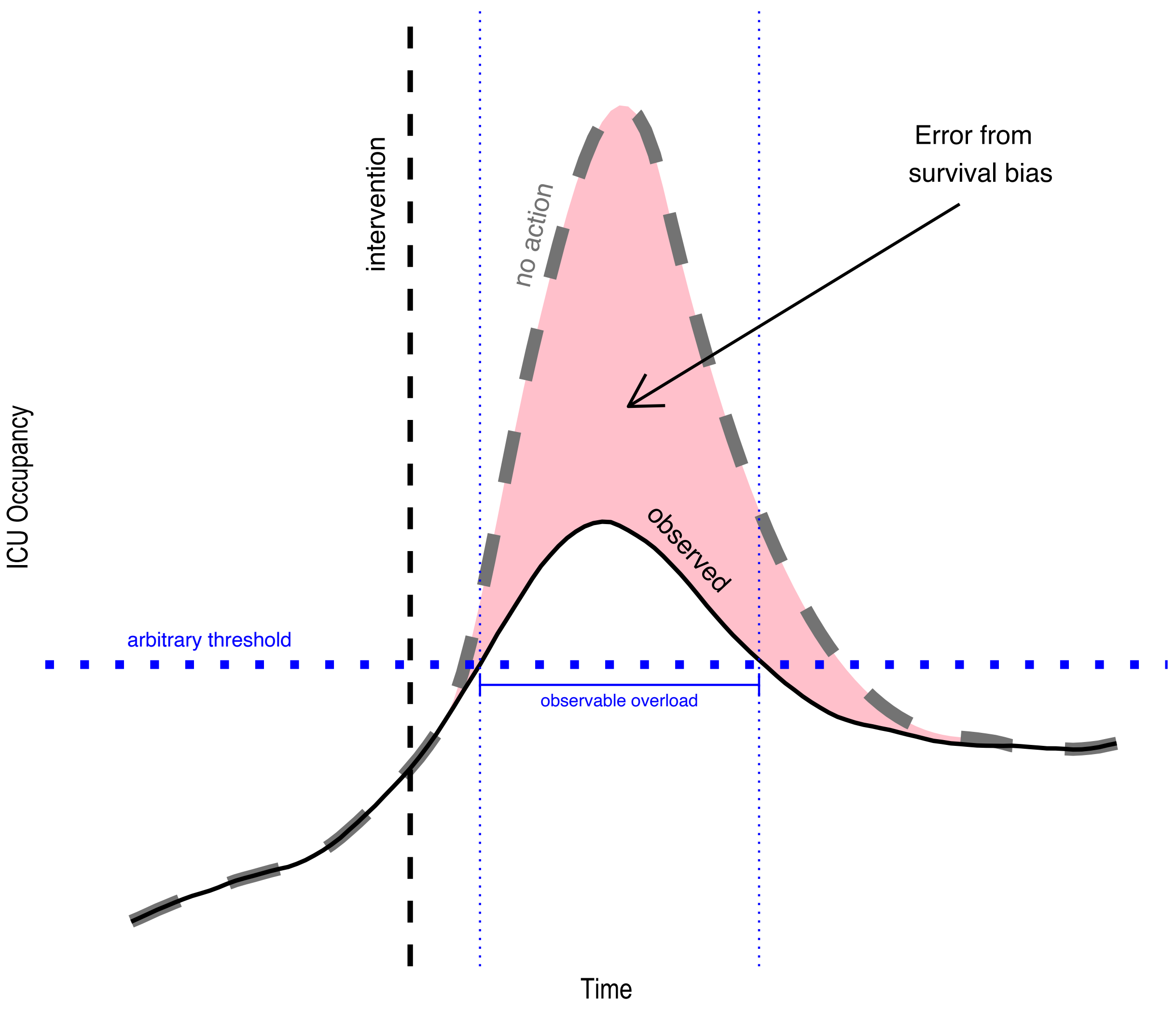

observed, is important to identify models that predict overload too often. We note that the same argument for robustness does not hold for areas of observable underload but emphasise that errors during these periods are not expected to be as severe.
